# Evidence for SARS-CoV-2 Delta and Omicron co-infections and recombination

**DOI:** 10.1101/2022.03.09.22272113

**Authors:** Alexandre Bolze, Tracy Basler, Simon White, Andrew Dei Rossi, Dana Wyman, Pavitra Roychoudhury, Alexander L. Greninger, Kathleen Hayashibara, Mark Beatty, Seema Shah, Sarah Stous, Eric Kil, Hang Dai, Tyler Cassens, Kevin Tsan, Jason Nguyen, Jimmy Ramirez, Scotty Carter, Elizabeth T. Cirulli, Kelly Schiabor Barrett, Nicole L. Washington, Pedro Belda-Ferre, Sharoni Jacobs, Efren Sandoval, David Becker, James T. Lu, Magnus Isaksson, William Lee, Shishi Luo

## Abstract

Between November 2021 and February 2022, SARS-CoV-2 Delta and Omicron variants co-circulated in the United States, allowing for co-infections and possible recombination events. We sequenced 29,719 positive samples during this period and analyzed the presence and fraction of reads supporting mutations specific to either the Delta or Omicron variant. We identified 18 co-infections, one of which displayed evidence of a low Delta-Omicron recombinant viral population. We also identified two independent cases of infection by a Delta-Omicron recombinant virus, where 100% of the viral RNA came from one clonal recombinant. In the three cases, the 5’-end of the viral genome was from the Delta genome, and the 3’-end from Omicron including the majority of the spike protein gene, though the breakpoints were different. Delta-Omicron recombinant viruses were rare, and there is currently no evidence that Delta-Omicron recombinant viruses are more transmissible between hosts compared to the circulating Omicron lineages.

## Introduction

Recombination is one way a virus can evolve to acquire a new combination of mutations. Recombinations have played an important role in the evolution of the RNA viruses HIV-1 and SARS-CoV-2 (Fischer et al., 2021; Jackson et al., 2021). In humans, the analysis of the first 87,695 SARS-CoV-2 genomes shared on GISAID in 2021 identified 225 sequences of likely recombinant origins (Varabyou et al., 2021). However, tracking recombinations for SARS-CoV-2 remains challenging because of the relatively low diversity of the genomes. Moreover, without the underlying sequencing data or orthogonal confirmation, it is difficult to determine whether recombinant sequences are real or due to contamination, technical artifacts or naturally occuring mutations shared by multiple variants.

The Omicron variant, first detected by scientists in Botswana and South Africa in early November 2021 (Viana et al., 2022), has rapidly spread across the globe. The variant includes the three Pango lineages BA.1 (Nextstrain clade 21K), BA.2 (Nextstrain clade 21L) and BA.3, and is characterized by a large number of mutations in the spike protein (Viana et al., 2022). In the United States, the first Omicron case was reported on December 1 (Vaziri and Allday, 2021), when the Delta variant was dominant (Bolze et al., 2021; Earnest et al., 2021). We hypothesized that during the subsequent period when Delta and Omicron co-circulated, some cases of co-infections would occur and potentially result in the emergence of a new SARS-CoV-2 variant resulting from the recombination of a Delta variant and an Omicron variant. This, in turn, would result in a new combination of mutations with unknown properties.

The aims of this study were (i) to look for cases co-infected with Delta and Omicron variants, and (ii) to study SARS-CoV-2 recombination in humans by analyzing co-infection samples, as well as looking for clonal infections caused by a Delta and Omicron recombinant.

## Results

### Co-circulation of Delta and Omicron variants in the United States

We sequenced and assigned a lineage to 29,719 samples positive for SARS-CoV-2 collected by the Helix laboratory across the United States (**Table S1**) between 22 November 2021 and 13 February 2022. These samples came from anterior nasal swabs of different individuals, with one viral sequence assay performed per person infected (similar to a cross-sectional analysis). The large majority of samples were collected at a national retail pharmacy. Samples from San Diego County were collected as part of community testing organized by San Diego County. The individuals tested represented a diverse range of race and age groupings (details in **Table S1**). We observed that the Omicron variant quickly grew to explain >99% of cases as of the week of January 17 (**Figure S1, Table S2)**. Delta and Omicron variants therefore co-circulated (each representing >1% of infections) from 6 December 2021 to 16 January 2022, represented by 14,214 sequences in our dataset (**Figure S1, Table S2**). During that time, the overall number of cases in the United States remained high, above 150,000 new cases per day and above a 7-day case rate of 250 per 100,000 individuals (CDC, 2020). The possibility of a co-infection by two distinct variants was therefore also high during this period.

### Co-infection with Delta and Omicron variants

When a person is infected by two distinct variants/viruses, multiple copies of the full genome of each variant are present in the sample. A fraction (*x*%) of the total extracted SARS-CoV-2 RNA will come from variant A, and the remaining fraction (100-*x*%) of the RNA will come from variant B. Sequencing at a high enough coverage will lead to calling mutations that define both variant A and variant B, but each mutation will only be supported by a fraction of the reads overlapping the given position; the mutations specific to variant A should be called with ∼*x*% of the reads overlapping the position, whereas the mutations specific to variant B should be called with (100-*x*)% **(Figure 1A**). In order to identify this co-infection signature, we selected a list of mutations specific to the Delta variant and a list of mutations specific to the Omicron variant (**Table S3**). All mutations selected had a call (not an “N”) in >95% of the samples between November 2021 and February 2022 (**Table S3**).

**Figure 1:**
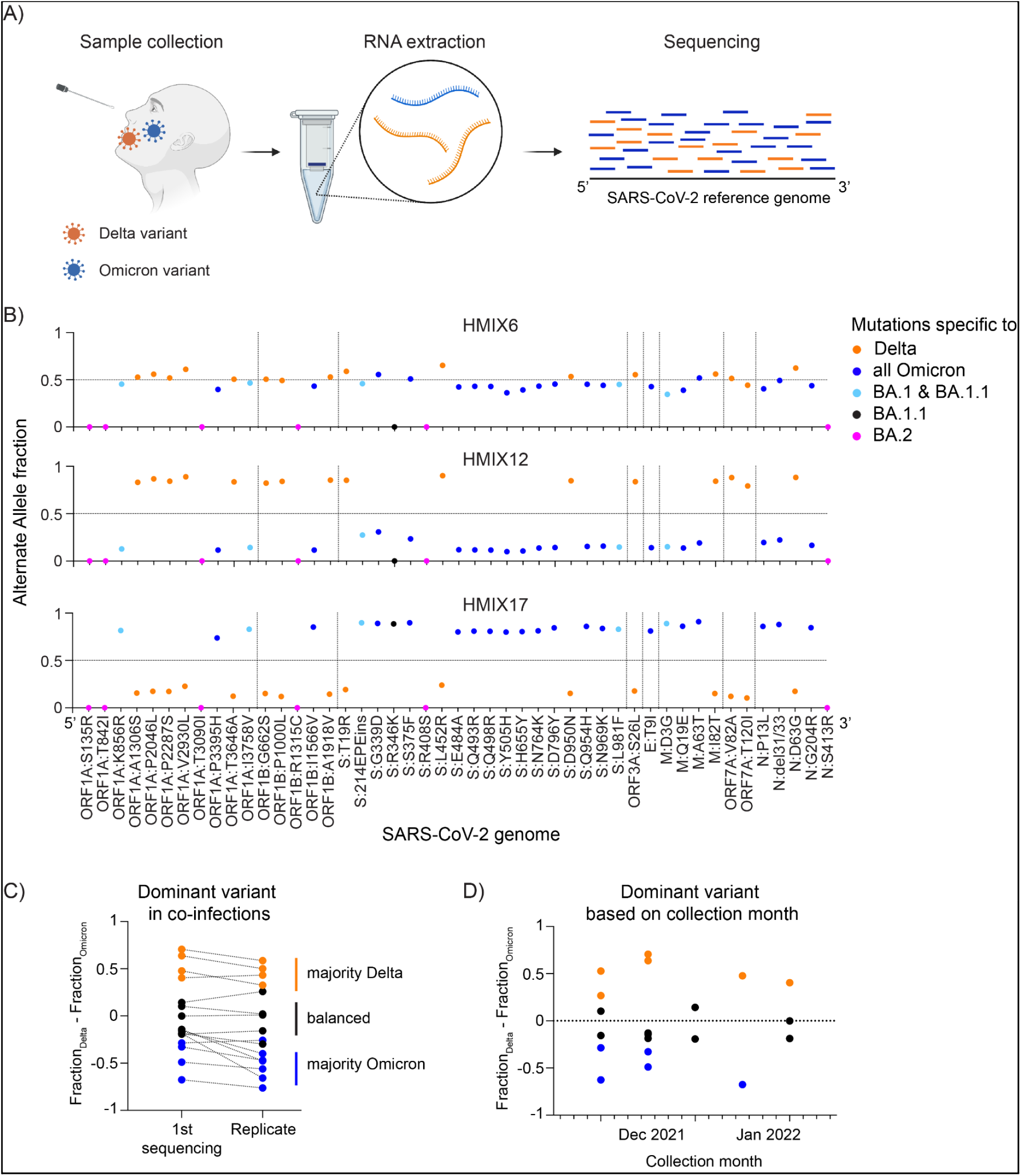
Co-infection with Delta and Omicron variants. (A) Schematic of a co-infection and the impact on the sequencing output. (B) Three example graphs representing the alternate allele fraction for each mutation. Forty-seven mutations are plotted in order of their position on the SARS-CoV-2 genome from 5’ to 3’. Genes are separated by dashed vertical lines. Sixteen mutations specific to Delta are represented in orange. Nineteen mutations specific to Omicron and shared by all its sub-lineages are represented in blue. Five mutations specific to BA.1 (and BA.1.1) are in light blue. The defining BA.1.1 mutation is in black and 6 mutations specific to BA.2 are in magenta. The graphs for all 18 co-infections are in **Figure S2**. (C) The difference of Delta fraction and Omicron fraction is plotted for the initial sample and the replicate sequenced after a new RNA extraction. Dashed lines connect identical samples. Orange dots represent samples with a majority of Delta. Blue dots represent samples with a majority of Omicron. (D) The difference of Delta fraction and Omicron fraction for the initial sample is plotted against the collection month of the sample. Orange dots represent samples with a majority of Delta. Blue dots represent samples with a majority of Omicron.

We identified 21 samples that were very likely to be co-infected with Delta and Omicron variants by filtering for samples where the median alternate allele fraction is less than 0.85 (see **Methods**). For 19 samples (two samples were unable to be re-tested), we validated the results by RNA re-extraction and re-sequencing. For a few samples, we also validated the results using an orthogonal genotyping assay (see **Methods**). The results replicated in 18 of the 19 samples (**Figure 1B, Figure S2, Table 1**), including two (HMIX1 and HMIX2) that have already been reported in a separate study (currently under review). A detailed list of the allele depths for each mutation for all of these samples is in **Table S4**. Overall, we estimate that on average ∼1 in 800 (18 / 14,214; 95% CI: 1/540 to 1/1,470, assuming a binomial distribution) positive samples between 6 December 2021 and 16 January 2022 had a co-infection.

**Table 1:** Samples with co-infections and Delta-Omicron infections.

| Name | Collection month | State | COVID Cq | Unique SARS-CoV-2 reads | Median AAF | Lineage | Clade | Replication method | Dominant variant | Breakpoint |
| --- | --- | --- | --- | --- | --- | --- | --- | --- | --- | --- |
| HMIX1 | Dec 2021 | CA | 18.1 | 480,979 | 0.71 | None | 21K (Omicron) | Re-extraction and re-sequencing | Omicron | Not Applicable |
| HMIX2 | Dec 2021 | NJ | 16.6 | 625,507 | 0.53 | None | 21M (Omicron) | Re-extraction and re-sequencing | balanced | Not Applicable |
| HMIX4 | Dec 2021 | CA | 19.5 | 826,167 | 0.61 | None | 21K (Omicron) | Re-extraction and re-sequencing | Omicron | Not Applicable |
| HMIX5 | Dec 2021 | PA | 19.2 | 1,164,229 | 0.54 | None | 21M (Omicron) | Re-extraction and re-sequencing | balanced | Not Applicable |
| HMIX6 | Dec 2021 | CA | 17.4 | 1,998,992 | 0.51 | None | 21M (Omicron) | Re-extraction and re-sequencing | balanced | Not Applicable |
| HMIX7 | Dec 2021 | PA | 19.3 | 320,813 | 0.43 | B.1.617.2 | 21J (Delta) | Re-extraction and re-sequencing | Delta | Not Applicable |
| HMIX8 | Dec 2021 | CA | 24.1 | 31,419 | 0.77 | BA.1 | 21K (Omicron) | Re-extraction and re-sequencing | Omicron | Not Applicable |
| HMIX10 | Dec 2021 | CA | 24.4 | 20,698 | 0.55 | None | 21K (Omicron) | Re-extraction and re-sequencing | Omicron | Not Applicable |
| HMIX11 | Dec 2021 | FL | 20.4 | 193,976 | 0.54 | None | 21K (Omicron) | Re-extraction and re-sequencing | balanced | Not Applicable |
| HMIX12 | Dec 2021 | GA | 21.1 | 359,378 | 0.83 | AY.25 | 21J (Delta) | Re-extraction and re-sequencing | Delta | Not Applicable |
| HMIX13 | Dec 2021 | MI | 21.1 | 668,533 | 0.51 | B.1.617.2 | 21J (Delta) | Re-extraction and re-sequencing | Delta | Not Applicable |
| HMIX14 | Dec 2021 | CA | 17.5 | 2,540,144 | 0.50 | None | 21M (Omicron) | Re-extraction and re-sequencing | balanced | Not Applicable |
| HMIX15 | Dec 2021 | TX | 22.9 | 41,983 | 0.51 | None | 21K (Omicron) | Re-extraction and re-sequencing | balanced | Not Applicable |
| HMIX16 | Jan 2022 | FL | 18.2 | 103,983 | 0.51 | None | 21J (Delta) | Re-extraction and re-sequencing | Delta | Not Applicable |
| HMIX17 | Jan 2022 | FL | 21.8 | 354,336 | 0.81 | BA.1.1 | 21K (Omicron) | Re-extraction and re-sequencing | Omicron | Not Applicable |
| HMIX18 | Jan 2022 | IN | 17.4 | 3,930,358 | 0.44 | None | 21M (Omicron) | Re-extraction and re-sequencing | Delta | Not Applicable |
| HMIX19 | Jan 2022 | OK | 19.9 | 1,121,368 | 0.54 | None | 21M (Omicron) | Re-extraction and re-sequencing | balanced | Not Applicable |
| HMIX20 | Jan 2022 | GA | 19.7 | 692,308 | 0.49 | None | 21M (Omicron) | Re-extraction and re-sequencing | balanced | Not Applicable |
| RECOMB1 | Jan 2022 | MA | 22.5 | 8,639 | 1.00 | None | 21K (Omicron) | Re-extraction and re-sequencing* | Not Applicable | 22,204 to 22,578 |
| RECOMB2 | Feb 2022 | MA | 20.5 | 240,742 | 1.00 | None | 21K (Omicron) | Re-extraction and re-sequencing | Not Applicable | 19,220 to 21,618 |

\*Recombination in this sample was also confirmed by an orthogonal genotyping array, and by long-read sequencing by an independent lab at the CDC (Lacek et al., 2022) .

Given how quickly Omicron displaced Delta, we hypothesized that in cases of co-infections we would see on average more Omicron virions compared to Delta. Using the fraction of sequencing reads that mapped to mutations in either Delta or Omicron as a proxy, the fraction of Delta and Omicron virions in a given sample appeared similar (between 40 and 60%) in 8 out of 18 co-infections (**Figure 1B, Figure S2, Table S5**). The Delta variant was higher than the Omicron variant in five co-infection samples, while the Omicron variant was higher than the Delta variant in the remaining five (**Figure 1B, Figure S2, Table S5**). The fraction of Delta and Omicron virions in each sample were similar in the replicates (**Figure 1C**) despite re-extraction. These results did not support the hypothesis that the Omicron variant would outcompete the Delta variant when in the same host. There was also no correlation between the period of co-infection and the dominant variant (**Figure 1D**), or between the viral load in the nose and the dominant variant (**Figure S3**). We were not able to test the hypothesis that vaccination or prior infection by SARS-CoV-2 would better control Delta and lead to a higher fraction of Omicron in these co-infected samples. Our analysis is also limited by the fact that we do not have information whether the exposure and seed infection by the two distinct variants happened at the same time, or if they followed each other.

### Within-host recombination of Delta and Omicron genomes

We hypothesized that a subset of host cells in a co-infection would inevitably contain both variants, and therefore have the potential to generate recombinants. If these recombinants were replication competent and replicated enough, then we would detect them in sequencing output, manifesting as a change in allele fraction of defining mutations near the recombination breakpoint.

Indeed, we find that HMIX16 (**Figure 2A**) exhibits precisely this characteristic. Alternative allele fractions for Delta mutations hover around 0.80 near the 5’ end of the genome, but drop to around 0.50 near the beginning of the S gene and remain at this level until the 3’ end of the genome. This profile suggests the presence of a Delta-Omicron recombinant with a breakpoint preceding the S:214EPEins. Upon examination of read-pairs sequenced from HMIX16 that spanned mutations unique for Delta and Omicron upstream of S:214EPEins, we found 4 read-pairs that supported a Delta-Omicron recombinant, 7 read-pairs that supported Delta only, and 10 that supported Omicron only (**Figure 2B**). The read pairs that supported a Delta-Omicron recombinant comprise the S:156/157del mutation of Delta on the 5’ end, and the S:212del of Omicron on the 3’ end. The existence of these three unique mutation profiles presents compelling evidence that a recombinant virus was generated during co-infection with a breakpoint region of 157 base pairs between positions 22,036 and position 22,193. We did not find read-pairs supporting Delta-Omicron recombination in the same interval in the other co-infection samples showing that these recombinations remain a rare event.

**Figure 2:**
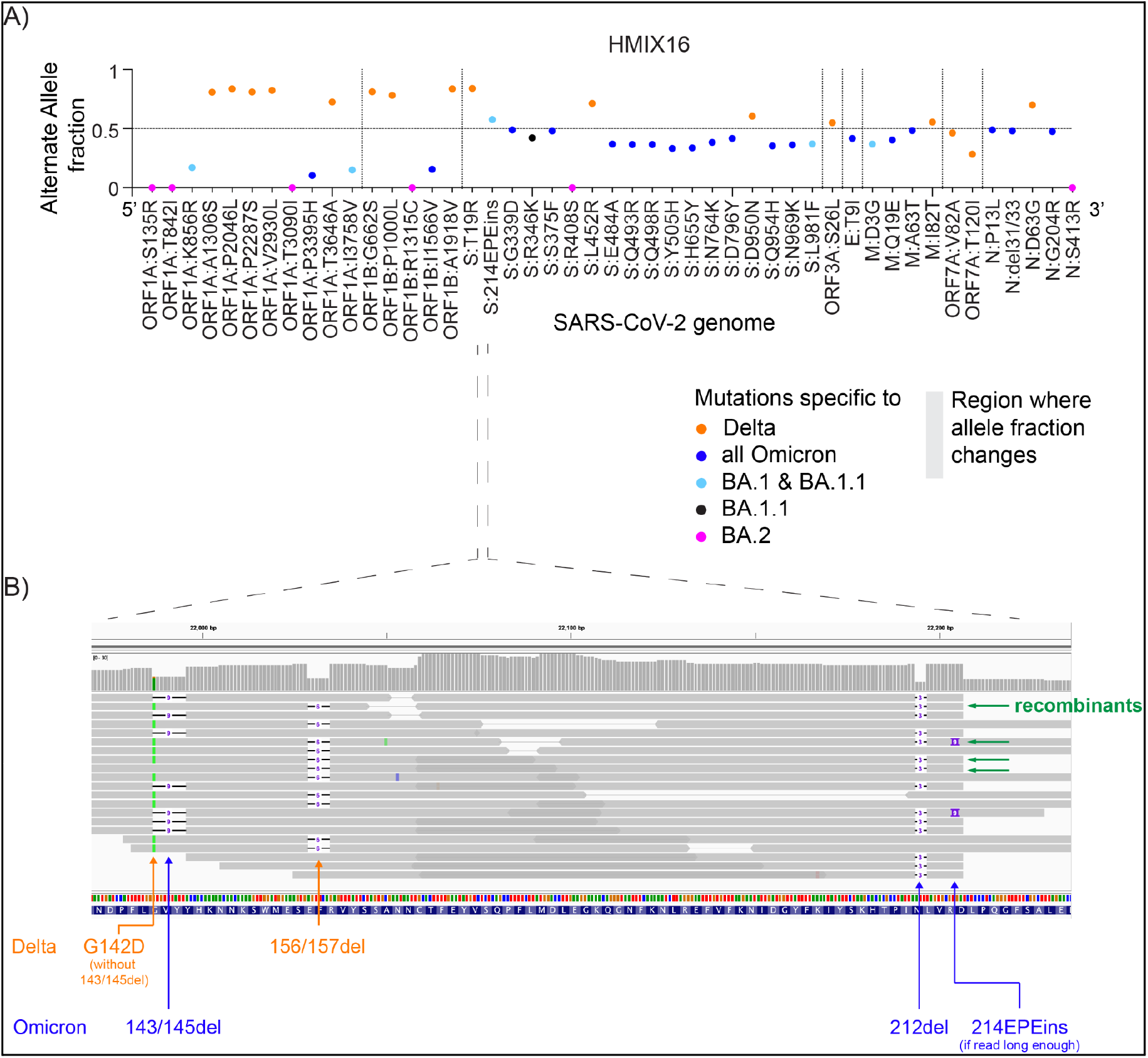
Evidence of Delta, Omicron, and recombinant read pairs in HMIX16. (A) Graph representing the Alternate Allele Fraction for each mutation of sample HMIX16. Forty-seven mutations are plotted in order of their position on the SARS-CoV-2 genome from 5’ to 3’. Genes are separated by dashed vertical lines. Sixteen mutations specific to Delta are represented in orange. Nineteen mutations specific to Omicron and shared by all its sub-lineages are represented in blue. Five mutations specific to BA.1 (and BA.1.1) are in light blue. One BA.1.1 mutation is in black and 6 mutations specific to BA.2 are in magenta. The gray box represents the region where the alternate allele fraction changes. (B) An IGV (Robinson et al., 2011) view of the alignments for HMIX16, subsampled to only include read pairs where the first in pair covers the S:156/157del position, and the second in pair covers the S:212del position. Read pairs representing three mutation profiles are present: (i) supporting Delta mutations only (7 read pairs), (ii) supporting Omicron mutations only (10 read pairs), and (iii) supporting a Delta/Omicron recombinant (4 read pairs) marked with a green arrow. Delta mutations in orange, Omicron mutations in blue. Read pairs that do not span these mutations are not shown.

### Infection with clonal Delta-Omicron recombinants

Having established that co-infections occur and can generate replication competent virus, we then looked for samples that are composed entirely of recombinant virus. In such samples, we expect that all mutations called would be supported by ∼100% of the reads because the viral population in the sample is composed of multiple copies of the same variant, rather than a mixture of two. We first looked for recombinants with one breakpoint where all mutations identified on the 5’-end of the breakpoint should be characteristic of one variant (e.g., variant A), and all mutations on the 3’-end of the breakpoint should be characteristic of the other variant (e.g., variant B) (**Figure 3A**). We identified seven samples that had Delta-specific ORF1A:A1306S at the 5’-end of the genome, and Omicron-specific N:P13L at the 3’-end. One sample had Omicron-specific ORF1A:P3395H at the 5’-end and Delta-specific N:D63G at the 3’-end. Further analysis of these eight genomes showed that only two genomes, RECOMB1 and RECOMB2, had multiple consecutive Delta mutations at the 5’-end while the 3’-end of the genome had all of the Omicron mutations but none of the Delta mutations (**Figure 3B**). Four of the six other genomes had all (5’ to 3’ of the genome) Omicron-specific mutations and the additional Delta ORF1A:A1306S, which was probably acquired independently. The remaining genomes had all of the Delta-specific mutations with one containing an additional Omicron N:P13L, and the other containing Omicron ORF1A:P3395H. These were probably also acquired independently.

**Figure 3:**
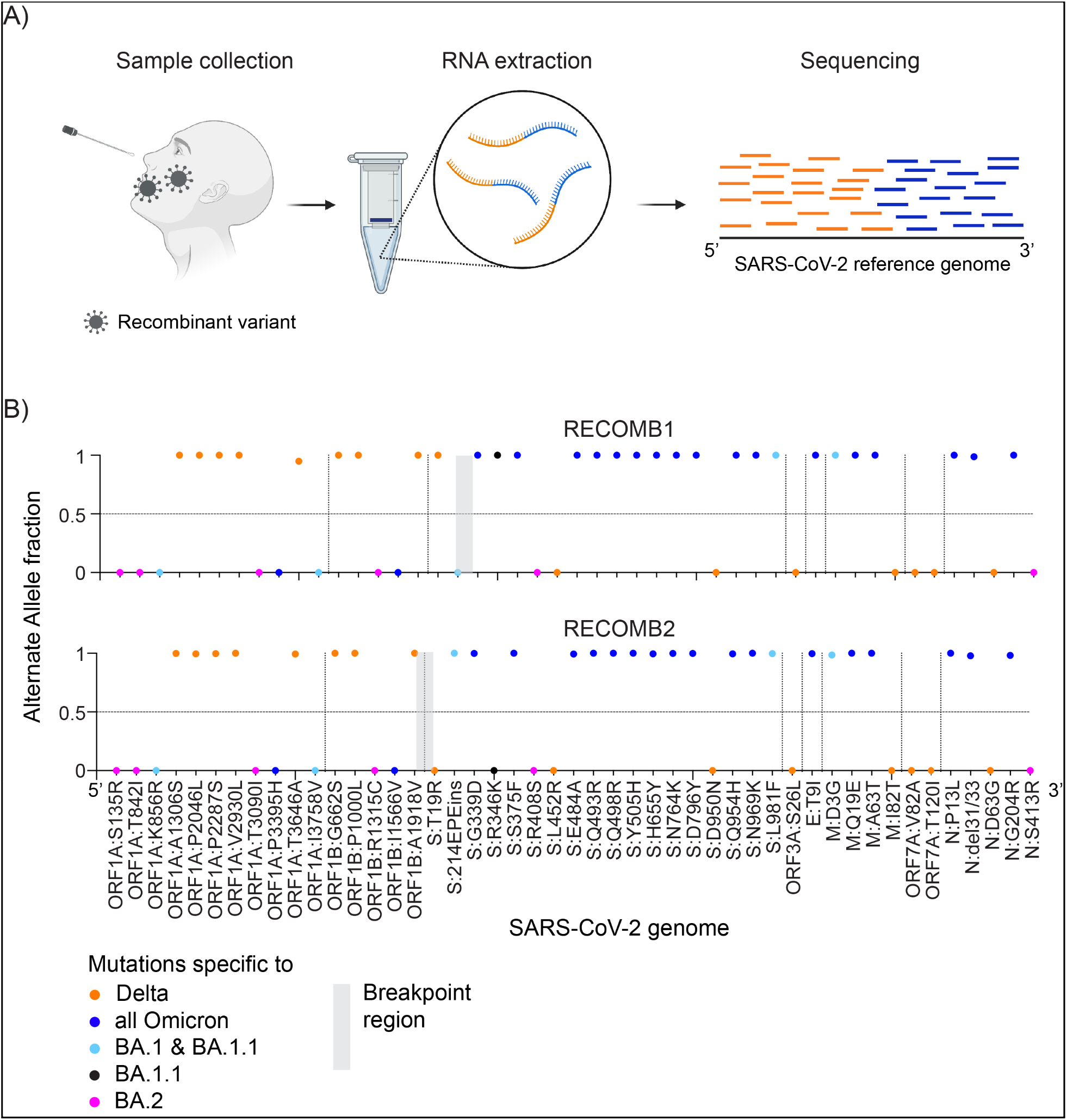
Infection with Delta-Omicron recombinants. (A) Schematic of the impact of an infection from a virus resulting from the recombination of Delta and Omicron on the sequencing output. (B) Graphs representing the alternate allele fraction for each mutation. Forty-seven mutations are plotted in order of their position on the SARS-CoV-2 genome from 5’ to 3’. Genes are separated by dashed vertical lines. Sixteen mutations specific to Delta are represented in orange. Nineteen mutations specific to Omicron and shared by all its sub-lineages are represented in blue. Five mutations specific to BA.1 (and BA.1.1) are in light blue. One BA.1.1 mutation is in black and 6 mutations specific to BA.2 are in magenta. The gray box represents the homologous region where the breakpoint of the recombination is.

Given that amplicon primer-based artifacts, in conjunction with laboratory contamination, have previously led to spurious signatures of recombination (Kreier, 2022), we were careful to check the quality of the two possible recombinant viruses. First, the library preparation method used by Helix’s viral sequencing protocol is hybridization-based capture, not amplicon. Hybrid capture is less susceptible to artifacts due to mutations at primer sites, which has been a recurring issue with possible recombinant viruses observed in GISAID (Sanderson and Barrett, 2021). We have also seen far less drop-out in sequences generated via hybrid capture compared to amplicon in our sequencing data (**Figure S4**). Second, the Cq values for these two infections were low Cq_(RECOMB1)_ = 22.5 and Cq_(RECOMB2)_ = 20.5, implying a high source viral load that would be less susceptible to contamination. Third, we replicated these results for RECOMB1 and RECOMB2 by re-extracting RNA from the collected sample and re-sequencing. In the initial and replicate samples, the median alternate allele fraction was 1, indicating that the majority of mutations were supported by 100% of the reads. Fourth, we performed a manual review of the alignments using IGV to make sure the reads supporting the mutations were of high quality. Fifth, we were able to validate our results for RECOMB1 by running a genotyping assay looking at Delta: C21618G (S:T19R at the protein level) and Omicron: G8393A, T13195C, C23202A (S:T547K at the protein level). The results showed the presence of Delta C21618G and Omicron C23202A in the sample, but the absence of Omicron: G8393A and T13195C. These confirm that the 5’-end of the genome was from Delta and the 3’-end from Omicron. Lastly, RECOMB1 sample was also sent to the US Centers for Disease Control and Prevention (CDC) laboratory and results were confirmed via another sequencing technology (Lacek et al., 2022). Together, these experiments provide evidence that the two independent infections were caused by viruses resulting from the recombination of Delta and Omicron.

The sequences of the two recombinant viruses differ slightly. The breakpoint region of RECOMB1 is 374 bases between position 22,204 and position 22,578, while the breakpoint region of RECOMB2 is 2,398 bases between position 19,220 and position 21,618 (**Figure 3B**). Of interest, there is a private mutation T19404C in RECOMB2 inside the breakpoint region. RECOMB1 is a recombination between Delta sublineage AY.119 and Omicron sublineage BA.1.1. The 5’ Delta end of RECOMB2 is too short for sublineage classification, but the 3’ end is Omicron sublineage BA.1. The full list of mutations including the presence of unlabeled mutations (not shared by a large fraction of genomes in the same lineage) are in **Table S6** (RECOMB1) and **Table S7** (RECOMB2) in a VCF-like format. These two samples were both collected in Massachusetts, but the difference in sequence suggests they are unrelated. Overall, infections from a recombinant Delta-Omicron virus remain rare: 2 out of 10,742 sequences between January 10 and February 13, 2022. Eight other sequences similar to RECOMB1 have been reported by the CDC from samples collected in the US from December 31, 2021 to February 12, 2022 (Lacek et al., 2022). We did not identify any recombinant with Omicron on the 5’-end and Delta on the 3’-end or any recombinant with two breakpoints in our dataset.

## Discussion

In this study we identified and validated 18 cases of co-infection with the Delta and Omicron variants, and 2 cases infected by a virus resulting from the recombination of Delta and Omicron. While contamination could lead to the same output as a co-infection, several pieces of evidence discount contamination: (i) re-extraction and re-sequencing these samples led to the same results; (ii) the fraction of reads supporting each variant was high in all cases (at least 15%); (iii) samples that showed a co-infection were collected and processed on different days, and other samples sequenced on the same plates did not show co-infection; (iv) the collection dates of the co-infections and the infections by a recombinant virus followed a logical timeline; (v) in one of these co-infections, we found evidence of recombinant virus at a low but detectable frequency, consistent with template-switching during viral replication in a cell infected with two variants.

In the case of the two recombinants, our data again supports chimeric sequences being the cause, rather than technical artifacts. First, we were able to replicate the result for both samples after re-extracting RNA. Second, our sequencing protocol is based on hybrid capture and is less prone to amplicon-based artifacts (**Figure S4**). Third, the chimeric exhibited breakpoint regions on the 5’ end of the spike protein as well as within the spike protein. These were not events limited to the spike protein, which is where many amplicon-based primer artifacts have been detected (Kreier, 2022).

Our study demonstrates the existence of co-infections, the presence of a recombinant population in at least one of these co-infections, and the existence of two infections consisting almost entirely of multiple copies of a recombinant virus. However, the mechanism by which a recombinant virus comes to dominate an infection remains somewhat of a puzzle. One possibility is that the two infections that contain only recombinant virus were themselves seeded by a recombinant virus. This implies that in their respective ancestral co-infections, the two recombinant viruses each rose to a high enough fraction to be transmitted during an exposure and were able to establish an infection in a new host. Yet, despite transmitting to a new host at least once, the transmission chain was not sustained; neither RECOMB1 nor RECOMB2 have led to large clusters of cases. The other possibility is that these two infections began as co-infections and that the recombinant viral population then completely outcompeted the Delta and Omicron populations within the host. Yet, it seems unlikely that Delta and Omicron can be completely cleared from a host, while leaving the recombinant virus population intact. In either case, the recombinant did not appear to have an increased ability to transmit between hosts compared to co-circulating Omicron variants. There are parallels here with HIV-1, where chronic infection and host immune response leads to extensive within-host diversity of the virus, but the genotypes of the virus that ultimately seed new infections are from a much narrower set of viral types (Joseph et al., 2015; Sagar et al., 2009).

With more diversity in circulating SARS-CoV-2 genomes, it will now be possible to track recombinations, characterize the rate of recombination, and identify hot spots for breakpoints (de Klerk et al., 2021). One way to detect these recombinants is the strategy we used. Another is to review every instance where a sample has good sequencing metrics but where methods like Nextclade (Hadfield et al., 2018) or Pangolearn (Rambaut et al., 2020) have difficulty attributing a clade or a lineage to the sequence. Specialized methods have also been developed to detect recombination in viruses (Martin et al., 2015; Samson et al., 2021; Varabyou et al., 2021). With a better understanding of SARS-CoV-2 recombination, and by drawing parallels with recombination in other unsegmented positive-strand RNA viruses, as well as other viruses in general (Simon-Loriere and Holmes, 2011), we can be better prepared to anticipate new variants or combinations of mutations of SARS-CoV-2 that may arise in the future.

## Methods

### Samples

This paper is based on the study of SARS-CoV-2 found and collected from individuals in the United States. The detailed demographics about these individuals can be found in **Table S1** of this paper.

The Helix data analyzed and presented here were obtained through IRB protocol WIRB#20203438, which grants a waiver of consent for a limited dataset for the purposes of public health under section 164.512(b) of the Privacy Rule (45 CFR § 164.512(b)). All samples were de-identified before receipt by the study investigators.

### Helix COVID-19 test data and sample selection

All viral samples in this investigation were collected by Helix through its diagnostic testing laboratory. The Helix COVID-19 test is run on specimens collected across the US, and results are obtained as part of our standard test processing workflow using specimens from anterior nares swabs. The Helix COVID-19 Test is based on the Thermo Fisher TaqPath COVID-19 Flu A, Flu B Combo Kit, which targets three respiratory pathogens (SARS-CoV-2, Influenza A, and Influenza B). Swabs are transported in saline and sample tubes are heat inactivated upon receipt at the lab. Test results from positive cases, together with a limited amount of metadata (including sample collection date, state, and qRT-PCR Cq values for all targets), were used to build the research database used here.

### SARS-CoV-2 sequencing and consensus sequence generation

Sequencing was performed by Helix as part of the SARS-CoV-2 genomic surveillance program in partnership with the Centers for Disease Control and Prevention (CDC). In the Helix workflow, RNA is extracted from 400 μL of patient anterior nares sample using the MagMAX Viral/Pathogen kit (ThermoScientific). Sequencing libraries were generated with total RNA library preparation (5μL RNA input volume) using the Rapid RNA Library Kit protocol (Swift Biosciences/Integrated DNA Technologies). SARS-CoV-2 genome capture was accomplished using hybridization kit xGen COVID-19 Capture Panel (Integrated DNA Technologies). Samples were sequenced using the NovaSeq 6000 Sequencing system S1 flow cell, with S1 Reagent Kit v1.5 (300 cycles). Note: Specimens identified as co-infections or recombinants through the surveillance program were verified by reprocessing from the original specimen. The process was replicated as described above; however, the hybridization probe panel was substituted. The IDT COVID-19 Capture Panel was replaced with the Respiratory Virus Research Panel (Twist Biosciences), while all other reagents remained the same.

Bioinformatic processing of this sequencing output is as follows. The flow cell output is demultiplexed with bcl2fastq (Illumina) into per-sample FASTQ sequences that are then run through the Helix fastagenerator pipeline to produce a sequence FASTA file. First, reads are aligned to a reference comprising the SARS-CoV-2 genome (NCBI accession NC_045512.2) and the human transcriptome (GENCODE v37) using BWA-MEM. Reads are then marked for duplicates before proceeding to variant calling using the Haplotyper algorithm (Sentieon, Inc). Finally, the per-base coverage from the alignment file (BAM) and per-variant allele depths from the variant call format (VCF) file are used to build a consensus sequence according to the following criteria: coverage from at least 5 unique reads is required with at least 80% of the reads supporting the allele. Otherwise, that base is considered uncertain, and an N is reported.

Alternate allele fraction is the number of reads supporting an alternate allele (i.e. a mutation) divided by the total number of reads covering the position. The median alternate allele fraction is calculated as the median value of alternate allele fractions at sites where at least 15% of the reads support a mutation.

### Viral lineage designation

Viral sequences were assigned a Pango lineage (Rambaut et al., 2020) using pangoLEARN (https://github.com/cov-lineages/pangoLEARN). For this analysis, pangoLEARN version 2022-02-02 with Pangolin software version 3.1.11 was used. We sequenced and were able to attribute a lineage to 29,719 sequences from samples collected between November 22, 2021 and February 13, 2022 for genomic surveillance purposes.

### Genotyping

The detailed genotyping method as well as the validation of the method used in this study are previously described (Lai et al., 2022). The four specific markers used were:

– Delta: C21618G
– Omicron: G8393A
– Omicron: T13195C
– Omicron: C23202A

### Relative fraction of each variant in co-infections

Number of RNA copies and coverage does vary across the SARS-CoV-2 genome. The density of mutations specific to Delta or Omicron also varies. There are many more Omicron-specific mutations in the spike protein. To try to minimize some biases, we took 4 Delta-specific mutations and 4 Omicron-specific mutations spread across SARS-CoV-2 genome to calculate the mean Delta-allele fraction and the mean Omicron-allele fraction in each sample. The 4 Delta-specific mutations are: ORF1A:P2046L, ORF1B:P1000L, S:T19R, M:I82T. The 4 Omicron-specific mutations are: ORF1A:P3395H, ORF1B:I1566V, S:N969K, M:A63T.

The results are in **Table S5**. We considered Delta to be the dominant variant if the Delta fraction was above 60% and the Omicron fraction was below 40%. We considered Omicron to be the dominant variant if the Omicron fraction was above 60% and the Delta fraction was below 40%. Other samples were considered balanced. To decide which variant was dominant for each sample for **Table 1**, we used the results of the initial sequencing.

### Statistics

The 95% Confidence Interval for the frequency of co-infections during co-circulation was calculated using the Python statsmodels.stats.proportion.proportion_confint package. `ci_low, ci_up = sm.stats.proportion_confint(18, 14214, alpha=0.05, method=‘normal’)`

## Data Availability

All data produced in the present work are contained in the manuscript.
SARS-CoV-2 sequences have been uploaded on GISAID.
BAM files have been uploaded to SRA Bioproject: PRJNA804575

## Data availability

All samples with a qc_status of pass were uploaded to GISAID.

GISAID identifiers for RECOMB1: hCoV-19/USA/MA-CDC-STM-HZEBR92XC/2022, EPI_ISL_9088187

GISAID identifiers for for RECOMB2: hCoV-19/USA/MA-CDC-STM-SP94WR2RW/2022, EPI_ISL_10114799

BAMs of co-infection samples and recombinant samples are available at SRA STUDY: PRJNA804575. Link: https://www.ncbi.nlm.nih.gov/Traces/study/?acc=PRJNA804575&o=acc_s%3Aa

SRA BioSample accessions:

HMIX16: SAMN26527328, bioproject: PRJNA804575

(https://www.ncbi.nlm.nih.gov/biosample/26527328)

RECOMB1: SAMN26527329, bioproject: PRJNA804575

(https://www.ncbi.nlm.nih.gov/biosample/26527329)

RECOMB2: SAMN26527330, bioproject: PRJNA804575

(https://www.ncbi.nlm.nih.gov/biosample/26527330)

## Acknowledgements

We thank the employees of Helix, members of the CDC SPHERES consortium and California CovidNET for discussions and help with logistics. We thank Elizabeth Ohlsen and Temet McMichael for discussions and help with the epidemiology of co-infections. We thank the healthcare workers, frontline workers, and patients who made the collection of this SARS-CoV-2 dataset possible. Special thanks to Anand Pai for helpful discussions on the parallels with HIV-1. We thank the NIH RADx initiative, which funded a portion of this work. This work has been supported by the Centers for Disease Control and funded in part by CDC Contract 75D30121C12730 (Helix).

## Supplementary Information

4 Supplementary figures and 7 supplementary tables

**Table S1**: Demographic information on the population where the samples were collected

**Table S2**: Counts and fraction of different lineages in the United States by week, related to Figure S1

**Table S3**: List of mutations used to identify and differentiate Delta and Omicron variants

**Table S4**: Reference and Alternate allele depth for each mutation for all HMIX (co-infections) and RECOMB (Delta Omicron recombinant) samples

**Table S5**: Fraction of Delta vs Omicron in each co-infection samples

**Table S6**: List of mutations identified in RECOMB1 sample

**Table S7**: List of mutations identified in RECOMB2 sample

**Figure S1:**
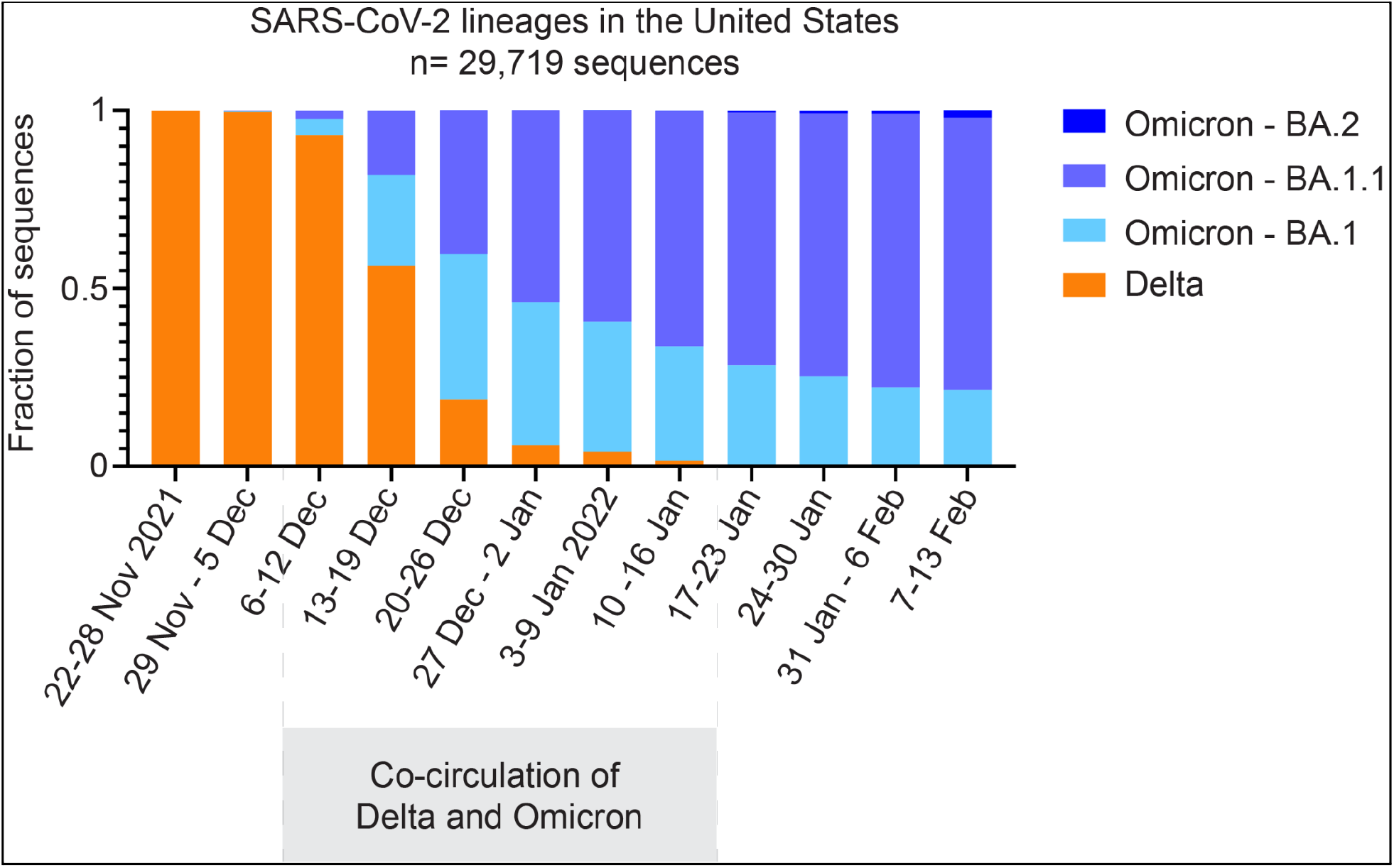
Co-circulation of Delta and Omicron variants in the United States. Fraction of lineages sequenced per week in the United States. Delta includes B.1.617.2 and all lineages starting with AY. Detailed numbers for the most common AY lineages are in **Table S2**. Delta: orange. Omicron: light blue (BA.1), blue (BA.1.1), navy blue (BA.2). The week of collection of the sample is on the X-axis.

**Figure S2:**
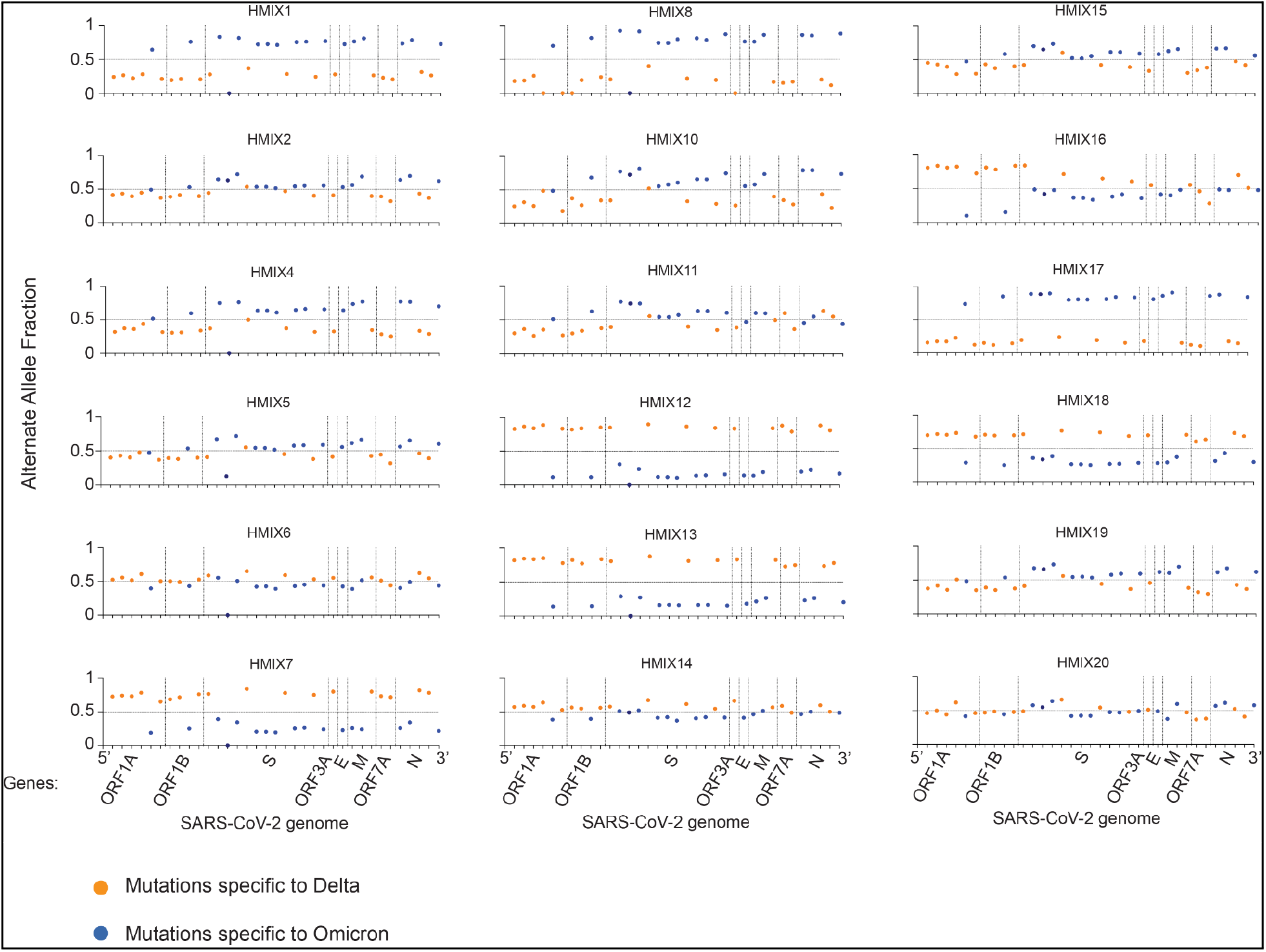
Co-infection with Delta and Omicron variants, related to Figure 1. Graphs representing the Alternate Allele Fraction for each mutation. 35 mutations are plotted in order of their position on the SARS-CoV-2 genome from 5’ to 3’. Genes are separated by dashed vertical lines. 18 mutations specific to Delta are represented in orange: ORF1A:A1306S, ORF1A:P2046L, ORF1A:P2287S, ORF1A:V2930L, ORF1A:T3646A, ORF1B:G662S, ORF1B:P1000L, ORF1B:1918V, S:T19R, S:L452R, S:P681R, S:D950N, ORF3A:S26L, M:I82T, ORF7A:V82A, ORF7A:T120I, N:D63G, N:R203M. 16 mutations specific to Omicron are represented in blue: ORF1A:P3395H, ORF1B:I1566V, S:G339D, S:S375F, S:E484A, S:Q498R, S:H655Y, S:N764K, S:D796Y, S:N969K, E:T9I, M:Q19E, M:A63T, N:P13L, N:DEL31-33, N:R203K. The S:R346K characteristic of Omicron BA.1.1 is in dark blue.

**Figure S3:**
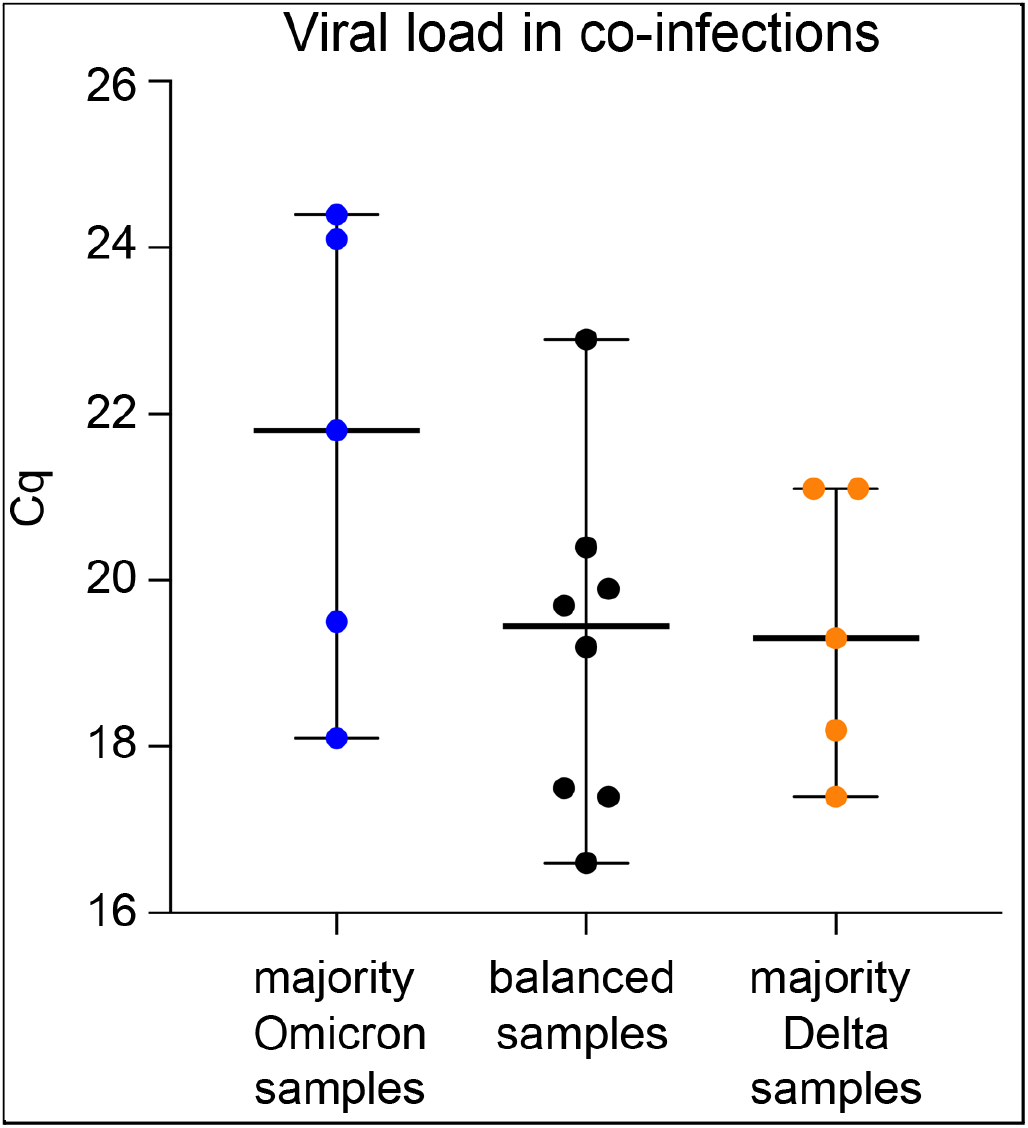
Viral load in co-infections. The Cq of each sample, grouped by the dominant variant. The first column are the samples (n=5) with a majority of Omicron; the second column are the samples (n=8) with a balanced mix of Delta and Omicron; the third column are the samples (n=5) with a majority of the Delta variant. Cq is based on the qRT-PCR assay performed on samples collected from the anterior nares of patients. Whiskers denote the median and 95% Confidence Interval. Of note, Omicron BA.1 and BA.1.1 samples have the S:69/70del, which interferes with one of the SARS-CoV-2 probes of the Thermo Fisher TaqPath COVID-19 Flu A, Flu B Combo Kit. Therefore 100% Omicron samples may be expected to have an elevated Cq compared to the Cq of a 100% Delta sample with the same viral load.

**Figure S4:**
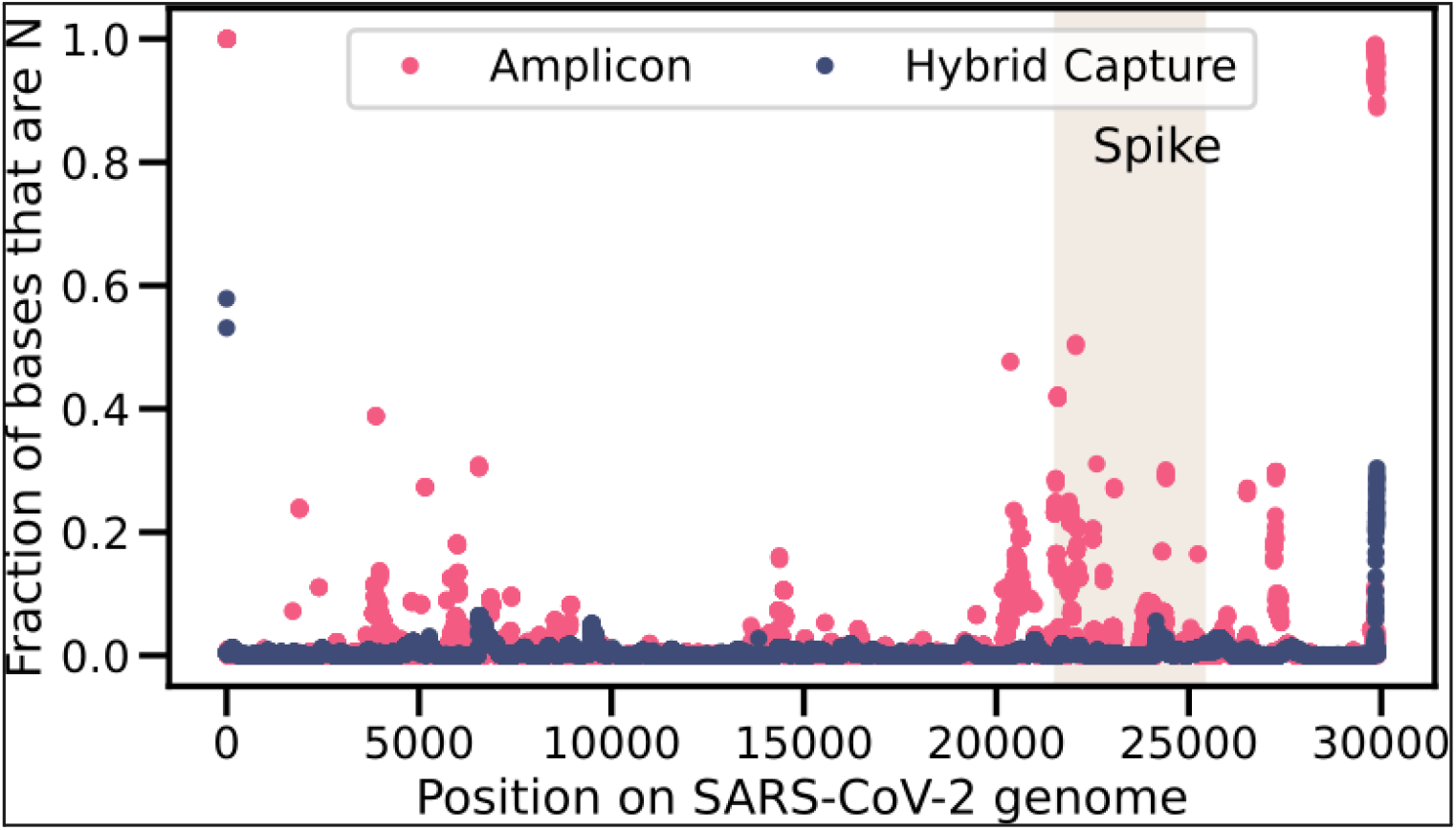
Comparison of sequencing output based on Amplicon vs. Hybrid capture. Fraction of sequences with an N by position in the SARS-CoV-2 genome. The shaded rectangle corresponds to the region that encodes the S glycoprotein. Fraction based on 2500 positive samples sequenced via the amplicon method and 2500 positive samples sequenced via hybrid capture. The 2500 samples sequenced via hybrid capture include 174 that were designated as Omicron. None of the 2500 samples sequenced via the amplicon method were designated Omicron.

